# Countries should aim to lower the reproduction number ℛ close to 1.0 for the short-term mitigation of COVID-19 outbreaks

**DOI:** 10.1101/2020.04.14.20065268

**Authors:** Michael E. Hochberg

## Abstract

The COVID-19 pandemic is still in its early stages and given the speed and magnitude of local outbreaks it is urgent to understand how mitigation measures translate into changes in key epidemiological and clinical outcomes. Here, we employ a mathematical model to explore the short-term consequences of lowering the reproduction number ℛ_0_ and delaying measures on total infections and fatalities. The positive implications of mitigation generally accrue as these measures are adopted early, with the most striking effects seen when the reproductive number is lowered to a level ℛ_C_≈1.0. As the delay in adopting measures exceeds approximately the half-way point to the peak of an outbreak, the effects of lowering ℛ_0_ markedly decrease. Aiming for reproduction numbers close to 1.0 can substantially reduce fatality probabilities over short time scales, particularly for larger populations. We conclude that research is urgently needed on how mitigation measures impact ℛ_0_ and how these can be optimized so as to achieve ℛ_C_≈1.0 whilst supporting individual freedoms, society and the economy.

## Introduction

As of April, 2020, COVID-19 is continuing to spread in many countries despite dedicated measures. Persistent spread could reflect multiple factors, including insufficient physical distancing or incomplete adoption of those measures, delays in outbreaks between areas within a given country, and inhomogeneities in local contact networks (*1,2*). Because of the explosive nature of new cases in the exponential growth phase of COVID-19, data may be incomplete and can lag behind their potential use in making important decisions for curbing outbreaks. Modeling analyses are central in this regard, since they can evaluate different scenarios to achieve the interlocked objectives of preserving health services, reducing individual morbidity and mortality, while maintaining social freedoms. Numerous promising avenues have been explored, and can be broadly characterized as conceptual tools for understanding outbreaks and for proposing control strategies, and more tactical instruments for actually applying strategies (e.g., *3-13*).

We previously simulated a simple SEIR model (*14*) to evaluate how short-term ‘bang-bang’ suppression-mitigation strategies could establish the conditions for a longer-term approach. A longer-term approach would integrate medicine (e.g., repurposed drugs, antibody transfusions, vaccines), technology (e.g., cell phone apps) and infrastructure (dedicated institutions) to reduce outbreaks and increase normalcy. The analysis showed that reacting insufficiently or increasingly late to an outbreak meant the need to enact more stringent ‘suppression’ measures to obtain a given level of reset in the numbers of infectious individuals. We found that this reset level, in turn, was crucial to how subsequent ‘mitigation’ measures could manage an outbreak. Importantly, approximate target levels were identified for lowered reproductive numbers, ℛ, that correspond to the rate of change in new cases.

More generally, a sequence of targets was proposed, based empirical observations of how countries have initially dealt with outbreaks, and a simple rational approach for future outbreak management. The sequence of targets that are being adopted – with each successive step following the perceived failure of the previous set of measures – take the symbolic form: (1) ℛ_0_ → (2) ℛ_C_<ℛ_0_ → (3) ℛ_C_<<1.0 → (4) ℛ_C_≈1.0 → (5) ℛ_C_<1.0, where ℛ_0_ is the basic reproductive number at the start of an outbreak (*15*) and ℛ_C_ is the level of ℛ that suppression or mitigation tactics strive to achieve in order to reduce the burden of an outbreak or meet specific objectives. Note that with the possible exception of a given country adopting or modifying measure packages engaged in other countries, governments *do not know* beforehand exactly what a given set of measures will achieve in terms of precise reductions in ℛ_0_ and the corresponding impacts on the outbreak. Many countries – for example, Italy, France, Spain, Germany and the United States – are currently in the second (suggested mitigation measures, e.g., social distancing, quarantining…) or third (enforced lockdowns) phase of this simple suite of strategies. A successful step 2 that actually results in ℛ_C_<1.0 may take the country directly to step 4, or possibly step 5, the latter which also seeks ℛ_C_<1.0, but does so through less socially burdensome measures, such as the introduction of repurposed drugs and vaccination.

We explore the short-term consequences of lowering ℛ_0_ (referred to hereafter as ‘mitigation’) and of delaying measures on the impacts of outbreaks in terms of the number of individuals eventually infected by the virus and fatalities. We find that the positive implications of mitigation accrue as these measures are adopted early, with the most striking effects as ℛ_C_→1.0. As the delay exceeds approximately the half-way point to the peak of an outbreak, the effects of lowering ℛ_C_ decrease. Finally, we show that lowering ℛ_C_→1.0 can substantially lower fatalities over fixed time scales, particularly for larger populations. We conclude that research is urgently needed on how measures impact ℛ_0_ and how these can be optimized so as to achieve ℛ_C_≈1.0, whilst supporting individual freedoms, society and the economy.

## Model

We employ a modified SEIR model of Susceptible (*S*) → Exposed (*E*) → Infectious (*I*) → Removed (*R*) states (*15*). The ordinary differential equations take the form:

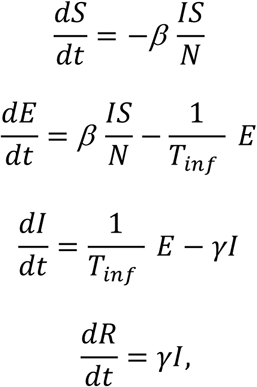

where *N* is a constant equal to *S+E+I+R, β* is the transmission parameter, *T*_*inf*_ is the infectious period, *γ* is the rate of removal into different subclasses of *R*, including mild cases, severve cases and fatalities. Specifically, here we focus on fatalities *C*_*F*_, given by

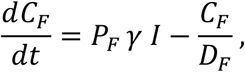

where *P*_*F*_ is the case fatality rate and *D*_*F*_ is the time from end of incubation to death (*T*_*itod*_) minus duration of infectiousness (*T*_*inf*_).

An outbreak occurs if the basic reproduction number, ℛ_0_ = *β / γ* > 1.0. The impact of control measures is easily understood by their impact on ℛ_0_, and in the presentation below, we refer to these effects by percent reductions in ℛ_0_, yielding the modified constant value, ℛ_C_.

## Numerical methods

The details of the numerical methods are described in (*14*). Briefly, the above model is an over-simplified representation of COVID-19 outbreaks. The purpose of its use is to generate first approximations of how mitigation efforts modeled by the proxy parameter ℛ_C_ could quantitively translate into the attenuation of outbreaks. Linking specific measures to their influence on key epidemiological parameters will be important in the future management of COVID-19.

The influences of reductions in ℛ_0_ on key epidemiological parameters were investigated using the Epidemic Calculator package (*16*) (Supplementary Material). This platform enables the user to set the parameters under focus here, namely, the total population size (*N*), the initial sub-population of infections individuals (*I*), the basic reproduction number (ℛ_0_), the day on which the mitigation measures begin and the resulting (lowered) reproduction number (ℛ_C_). The platform produces output in a display that the user can consult by clicking on the day or period of interest (**Figure 1**). As discussed in *14*, the accuracy of the Epidemic Calculator package has not been extensively tested, and the platform has several shortcomings. Caution is therefore necessary in interpreting the results presented below.

**Figure 1.**
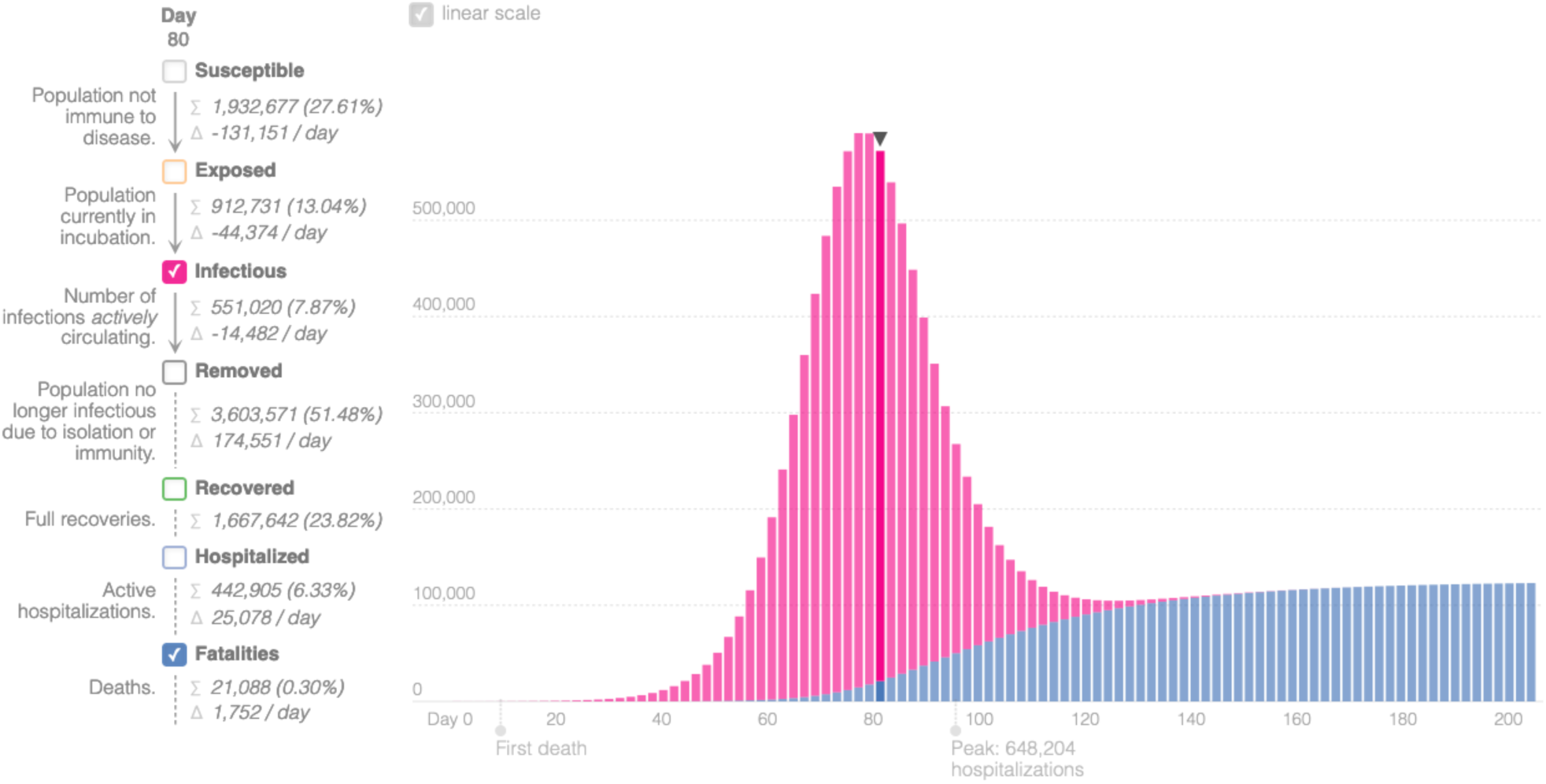
Visualization of output from the Epidemic Calculator (*16*), showing daily numbers of currently infectious individuals and cumulative fatalities in a population of 7 million. Statistics for Day 80 shown on the left. Initial number of infectious individuals=100. Other parameter values in Table 1.

The model parameter values employed in this study are presented in **Table 1**. Estimates of ℛ_0_ for COVID-19 vary approximately four-fold from *c*.1.5 to 6.5 (*17-20*), and we adopt a modal value of 2.5. Although the quantitative model predictions differ for lower and higher values of ℛ_0_ in the observed range, the qualitative patterns remain the same.

**Table 1.** Parameters and their baseline values employed in this study. See (*16*) and main text for details on parameter value sources.

| Parameter | Value |
| --- | --- |
| Basic Reproduction Number ($\mathcal{R}_0$) | 2.5 |
| Removal rate ($\gamma$) | 1/5.2 days |
| Duration patient is infectious ($T_{inf}$) | 2.9 days |
| Case fatality rate ($P_F$) | 0.02 |
| Time from end of incubation to death ($T_{itod}$) | 32 days |

## Results

The fraction of the population infected by SARS-CoV-2 depends on the start day of mitigation measures, the time horizon, and the impact of measures on ℛ_0_ (**Figure 2**). As expected, delaying intervention results in larger fractions of a population infected at any given time, and infected levels are lower for the shorter (80 day) compared to the longer (200 days) time horizons. However, the differences between the two time-horizons decrease as impacts on ℛ_0_ increase (*cf*. values beyond a 60% decrease in ℛ_0_ – corresponding to ℛ_C_≲1.0 – in **Figs. 2A,B**). Note too that sensitivity to start date increases as the time horizon decreases, but this effect is most appreciable for intermediate reductions in ℛ_0_ (e.g., *cf*. 20% to 40% reduction in **Figs. 2A,B**). In other words, whereas achieving 1.5≲ ℛ_C_<ℛ_0_ – if commenced early – can result in a substantial reduction in total infections by day 80 (**Fig. 2A**), the effect is largely erased on the longer time horizon of 200 days (**Fig. 2B**). Interestingly, small delays in starting mitigation measures from the start of a simulation may make little difference to the total number eventually infected. This effect is most noticeable between start days 10 and 30, as mitigation exceeds a 60% or greater decrease in ℛ_0_ (i.e., ℛ_C_≲1.0), and for the longer time window of 200 days. Note that the lack of sensitivity between start days for lower mitigation in **Fig. 2B** can be explained by the fact that the epidemic is virtually complete by 200 (see **Fig. 1**). These results thus underscore how attaining mitigation objectives is contingent on the time frame, with rapid responses having short term benefits for a range of mitigation levels (**Fig. 2A**), whereas they have disproportionately higher benefits in the longer-term only for sufficiently pronounced mitigation levels (**Fig. 2B**).

**Figure 2.**
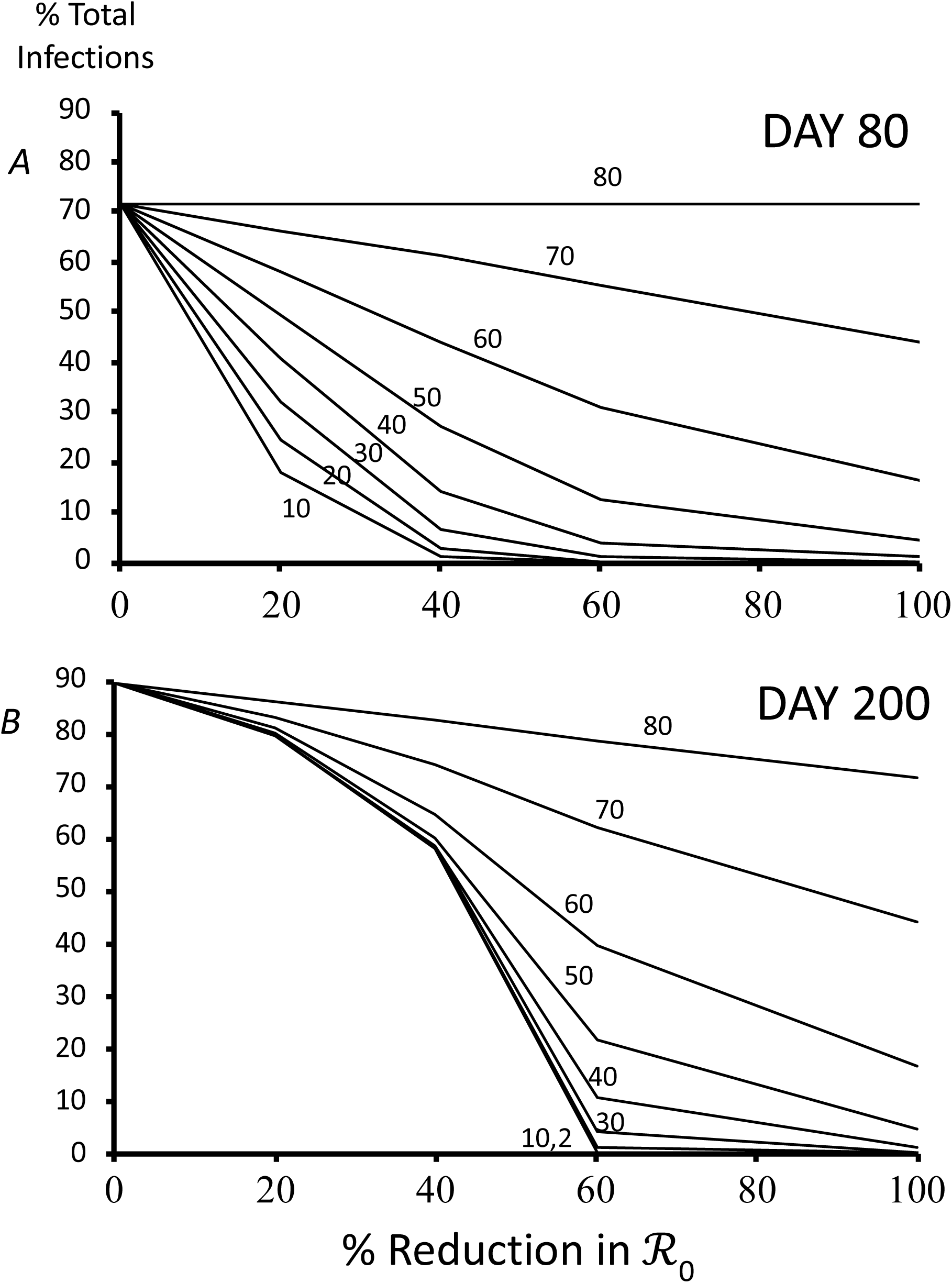
Effect of mitigation measures (represented as % reductions in ℛ_0_), number of days elapsed before mitigation begins (numbers next to lines), and number of days of the simulation (*A* 80 days; *B* 200 days), on the percentage of the population infected. At ℛ_0_, we observed 72% of the population had been infected at 80 days and 89% at 200 days. Lines connecting points at 0, 20, 40, 60 and 100% reduction in ℛ_0_ aid visualization. Assume population of 7 million inhabitants and 100 initial number infectious cases. Other parameters as in Table 1.

Lives saved by mitigation measures is influenced by the same factors considered above, but in an inverse way compared to total infections (*cf*. **Figs. 2,3**). Notably, reductions in ℛ_0_ approaching and exceeding 60%, corresponding to ℛ_C_≲1.0, have major benefits to reducing fatalities, particularly when measures are started with minimum delays (**Fig. 3**). Delays of 40 or more days never exceed saving more than 50% of lives that would have been lost without measures, even if the reproduction number is lowered by 100% to 0.0. As for the total number of individuals infected, the shortcomings of insufficient impacts on ℛ_0_ accrue with time (*cf*. 20%-40% reductions in **Figs. 3A,B**).

**Figure 3.**
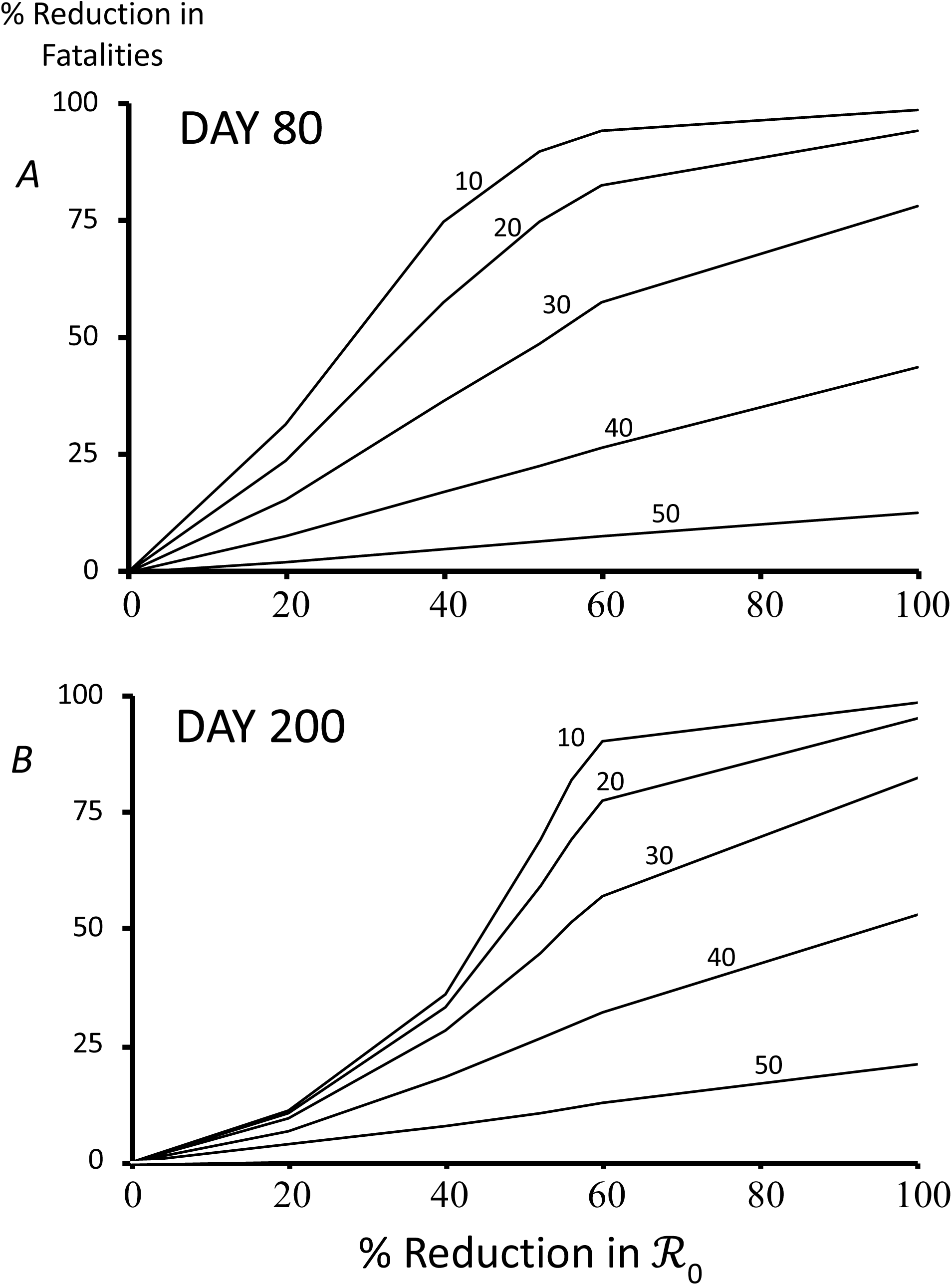
Effect of mitigation measures (represented as % reductions in ℛ_0_), number of days elapsed before mitigation begins (numbers next to lines), and number of days of the simulation (*A* 80 days; *B* 200 days), on the percentage of fatalities averted compared to no mitigation measures. Assume population of 10 million and 10,000 initial infectious individuals. At ℛ_0_, we observed 111,437 deaths at 80 days and 177,756 at 200 days. Lines connecting points at 0, 20, 40, 60 and 100% reduction in ℛ_0_ aid visualization. Other parameters as in Table 1.

Our previous work suggested that population size was important in the impact of mitigation measures (*14*). We find that the fraction of lives saved in a given time frame can be strongly associated with population size (**Figure 4**). The sensitivity in fatality reduction is maximal intermediate reductions in ℛ_0_. For example, an individual who would have died in the first 80 days without mitigation measures has a virtually 100% chance of surviving based on mitigation measures that reduce ℛ_0_ by 40% or more, and if living in a population with *c*.1 billion inhabitants. In contrast, survival chances drop below 50% under the same conditions in a population of *c*.100,000 individuals. Similar to the observations in the previous paragraphs, longer mitigation time frames require stronger impacts on ℛ_0_ to achieve equivalent benefits of shorter ones in terms of the percentage of lives saved (*cf*. **Figs. 4A,B**).

**Figure 4.**
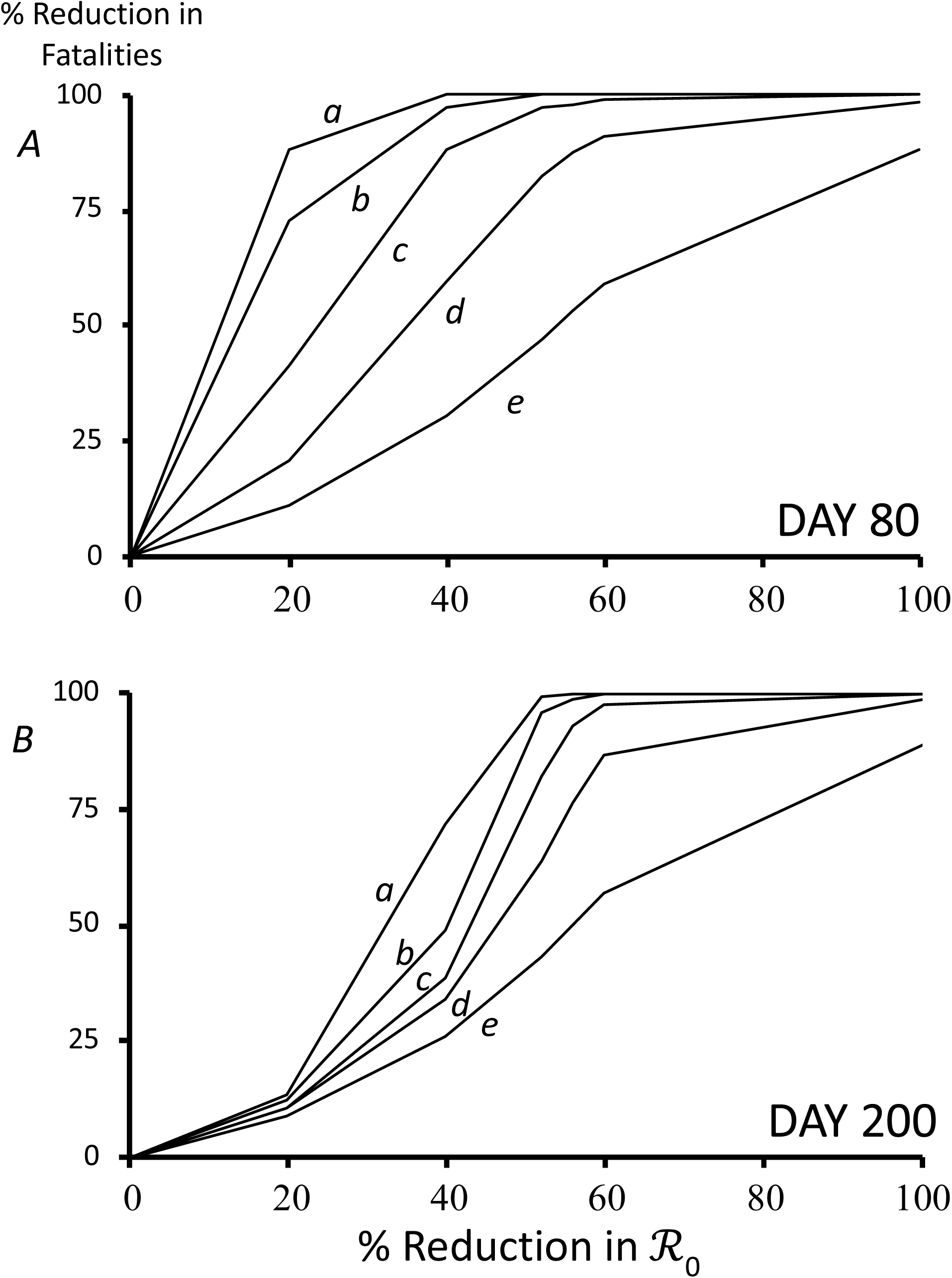
Effect of mitigation measures (represented as % reductions in ℛ_0_), population size (letters next to lines), and number of days of the simulation (*A* 80 days; *B* 200 days), on the percentage of fatalities averted compared to no mitigation measures. *a=*1 billion, *b*=100 million, *c*=10 million, *d*=1 million, *e*=100 thousand individuals. Assume 10,000 initial infectious individuals. At ℛ_0_, we observed 111,437 deaths at 80 days and 177,756 at 200 days. Lines connecting points at 0, 20, 40, 60 and 100% reduction in ℛ_0_ aid visualization. Other parameters as in Table 1.

## Conclusions

The results support the patterns observed in our previous study showing how lowering ℛ_0_ to approximately 1.0 is a sensible strategy for mitigating outbreaks on time scales of several weeks to several months (*14*). Measures falling too short of the ℛ_C_≈1.0 target can mean substantially diminished outcomes and the likelihood that additional measures are needed to mitigate the outbreak. This is in effect what we have observed in different countries, where sensible, but untested measures such as physical distancing, travel restrictions, washing hands frequently, coughing or sneezing away from others, and self-quarantining were insufficient in stemming new cases on time scales of days to weeks (*21*). On the other hand, further reductions below ℛ_C_≈1.0 may have specific objectives (e.g., steps 3 and 5 in the Introduction section), but come with increased constraints to personal freedoms and costs to the economy.

Our previous work suggested that population size was an important parameter in how an outbreak unfolds and the impact of mitigation measures (*14*). We found that larger populations benefit more in terms of percentage reduced fatalities than do smaller populations. This can be explained in part because the same initial number (10000) of infectious individuals was used in all simulations, meaning that smaller countries were effectively further along in their outbreaks than were more populous ones. Nonetheless, this result and the findings of our previous study indicate that – all else being equal – larger populations have more time to reduce morbidity and mortality in outbreaks than do smaller ones. This insight could apply either to the homogenous populations assumed here, or to spatially subdivided ones, where we would expect delays in outbreaks between different localities. Further investigation is needed to establish how country size may correlate with other potentially important epidemic parameters such as population density and the age and spatial structure of contact networks.

We conclude that the key short-term target of mitigation measures is to lower the reproductive number as close as possible to 1.0, but that values slightly above are acceptable, especially if the numbers of circulating cases is very low. Mitigation measures engaged once an outbreak approaches its peak will have substantially less impact on infections and lessened fatalities than when started early. Given that different combinations of measures and their detailed deployments will influence ℛ, empirical and modeling research is urgently needed to establish optimal packages that prioritize those measures also sustaining individual freedoms, society and the economy.

## Data Availability

Epidemic simulator code provided in Supp Material

## Acknowledgements

I thank the members of the EEC research team at the Institute for Evolutionary Sciences and Joshua Weitz for discussions.

## Notes

### Competing Interest Statement

The authors have declared no competing interest.

### Funding Statement

No external funding received

